# A global overview of genetically interpretable comorbidities among common diseases in UK Biobank

**DOI:** 10.1101/2021.01.15.21249242

**Authors:** Guiying Dong, Jianfeng Feng, Fengzhu Sun, Jingqi Chen, Xing-Ming Zhao

## Abstract

**Background:** Comorbidities greatly increase global health burdens, but the landscapes of their genetic factors have not been systematically investigated.

**Methods:** We used the hospital inpatient data of 385,335 patients in UK Biobank to investigate the comorbid relations among 439 common diseases. Post-GWAS analyses were performed to identify comorbidity shared genetic risks at the genomic loci, network, as well as overall genetic architecture levels. We conducted network decomposition for interpretable comorbidity networks to detect the hub diseases and the involved molecules in comorbidity modules.

**Results:** 11,285 comorbidities among 439 common diseases were identified, and 46% of them were genetically interpretable at the loci, network, or overall genetic architecture level. The comorbidities affecting the same and different physiological systems showed different patterns at the shared genetic components, with the former more likely to share loci-level genetic components while the latter more likely to share network-level genetic components. Moreover, both the loci- and network-level genetic components shared by comorbidities mainly converged on cell immunity, protein metabolism, and gene silencing. Furthermore, we found that the genetically interpretable comorbidities tend to form network modules, mediated by hub diseases and featuring physiological categories. Finally, we showcased how hub diseases mediating the comorbidity modules could help provide useful insights into the genetic contributors for comorbiditities.

**Conclusions:** Our results provide a systematic resource for understanding the genetic predispositions of comorbidity, and indicate that hub diseases and converged molecules and functions may be the key for treating comorbidity. We have created an online database to facilitate researchers and physicians to browse, search or download these comorbidities (https://comorbidity.comp-sysbio.org).

## Background

Comorbidity, the coexistence of more than one disease in a patient not by chance, presents great challenges for disease diagnosis and treatment [1, 2]. Compared with single diseases, comorbidities are usually associated with more adverse health outcomes, such as lower quality of life and higher mortality rate, and with higher economic burden [3-5]. Understanding the mechanisms of comorbidities may be helpful for their early diagnosis, treatment and management, thereby helping reduce the global disease burden associated with comorbidity.

During the last decade, large-scale genome-wide association studies (GWASs) have found overlapped genetic risks for a few frequently comorbid diseases at the genomic loci level, i.e. single-nucleotide polymorphisms (SNPs) or genes, suggesting that there might be a molecular basis of comorbid relations. For example, GWASs have uncovered 38 SNPs associated with both asthma and allergic diseases [6], and 244 genome loci associated with ankylosing spondylitis, Crohn’s disease, psoriasis, primary sclerosing cholangitis and ulcerative colitis [7]. Additionally, Sánchez-Valle *et al*. found that disease interactions inferred from similarities between patients’ gene expression profiles have significant overlaps with epidemiologically documented comorbid relations [8], further supporting the genetic basis of comorbidity. Moreover, several studies also point out that diseases with high probability of concurrency tend to share more genes [9, 10]. These findings have added useful information to inform the biological etiology of comorbidity [10, 11].

The malfunctions caused by disease risk loci can spread via cellular networks owing to molecular interactions among genes. To this end, some studies capture the genetic overlaps between comorbidities by network level evidence, including protein-protein interactions (PPIs) and molecular pathways [9, 12, 13]. For example, Park *et al*. found a significantly positive correlation between the number of shared PPIs and the extent of disease concurrency by combining information on cellular interactions, disease–gene associations, and Medicare data [9]. Moreover, a significantly increased number of shared pathways between cancers and comorbid Mendelian diseases have also been observed [10]. These results indicate that dysfunctional entanglement in molecular networks might contribute to the coexistence of comorbid diseases in a patient. Additionally, some comorbidities have also been reported to be similar in overall genetic architectures measured by genetic correlations, such as the widespread genetic correlations among comorbid psychiatric disorders [14-17].

Due to the limited access to the matched epidemiological and genomic data of the same population group, existing studies either used matched data for a limited number of diseases, or collected large-scale genomic data and epidemiology data from different sources. However, the separation of genetic and epidemiological data makes it tricky to decide whether the shared genetic risks identified from one group can actually explain comorbidity identified in another group. In the past few years, the UK Biobank (UKB) has collected hospital inpatient data and genetic data for about four hundred thousand individuals, providing a unique opportunity to investigate the genetics underlying comorbid relationships between hundreds of common diseases [18].

In this study, we take advantage of the large scale, matched epidemiological and genetic data hosted in UKB to systematically investigate the comorbid relationships among 439 common diseases as well as their shared genetic factors. We have identified the comorbidity shared genetic components at loci and network levels, and performed the functional analyses on them to uncover converged biological functions. Furthermore, the shared genetic patterns of the comorbidities affecting the same and different physiological systems have been explored. Finally, we construct and decompose two comorbidity networks to find the hub diseases that mediate lots of comorbid relationships in comorbidity modules and to highlight the corresponding molecular mechanisms. Our results provide a systematic resource of comorbidities among common diseases, as well as their shared genetic risks (online database at https://comorbidity.comp-sysbio.org). The converged biological molecules and functions identified in this study are responsible for many comorbidities, which may serve as the key factors for management and treatment of comorbidity.

## Methods

### Population data

Population data used in this study is collected from the UKB [18]. More than 500,000 individuals aged 40-69 living in the UK were recruited to the assessment centers and signed an electronic consent to allow a broad range of access to their anonymized data for health-related research.

### Disease selection and classification

In UKB, field-41270 is the summary diagnoses for 410,293 patients across all their hospital inpatient records, which are coded according to the International Classification of Disease version 10 (ICD10). A total number of 11,727 ICD10 codes are recorded with affected patients. We define common diseases as the level 2 ICD10 codes of chapter I ∼ XIV with prevalence > 0.1%, since most of the publically available GWAS summary statistics of the UKB diagnoses are based on level 2 codes [19, 20]. Patients diagnosed with codes at or under level 2 are considered as suffering from the corresponding level 2 diseases. Due to the too-detailed phenotype description of ICD10 codes, such as F20 Schizophrenia and F25 Schizoaffective disorders, we further aggregated the highly similar ICD10 codes into one disease according to the phecodes [21]. Phecode is a collection of manually curated phenotypes by experts with the advantage of better aligning with diseases mentioned in clinical and genomic research. Finally, the diseases are manually classified, mostly according to their affected physiological system while also considering their origins [22].

## Results

### Comorbidities among common diseases in UKB

439 common diseases (prevalence > 0.1%) are selected for comorbidity analysis from UKB hospital inpatient data (see Methods for details, Additional file 2: Table S1), covering 385,335 patients. The average age of the patients for their first hospital diagnosis is 54, with more female patients than male patients (55% VS. 45%).

In all, 11,285 comorbidities are identified, involving 438 out of the 439 diseases, with D21 (“other benign neoplasms of connective and other soft tissue”) being the only exception (RR >1, *P*-value < 4.1e-6 with Bonferroni correction for all disease-pairs with RR > 1, Fig. 1a, Addiitonal file 2: Table S2; see Methods for details). Most diseases have less than 100 comorbid partners (average of 51, Fig. 1b). We observe that diseases with high prevalences tend to have more comorbid partners (Pearson’s correlation, *r* = 0.69, *P*-value = 3.3e-64; Additional file 1: Fig. S3). For example, the top three diseases with the most comorbid partners--Hypertension (I10), Hyperlipidemia (E78), and Type 2 diabetes (E11), have a prevalence of 27.5%, 13.1%, and 7.1%, respectively.

**Fig. 1.**
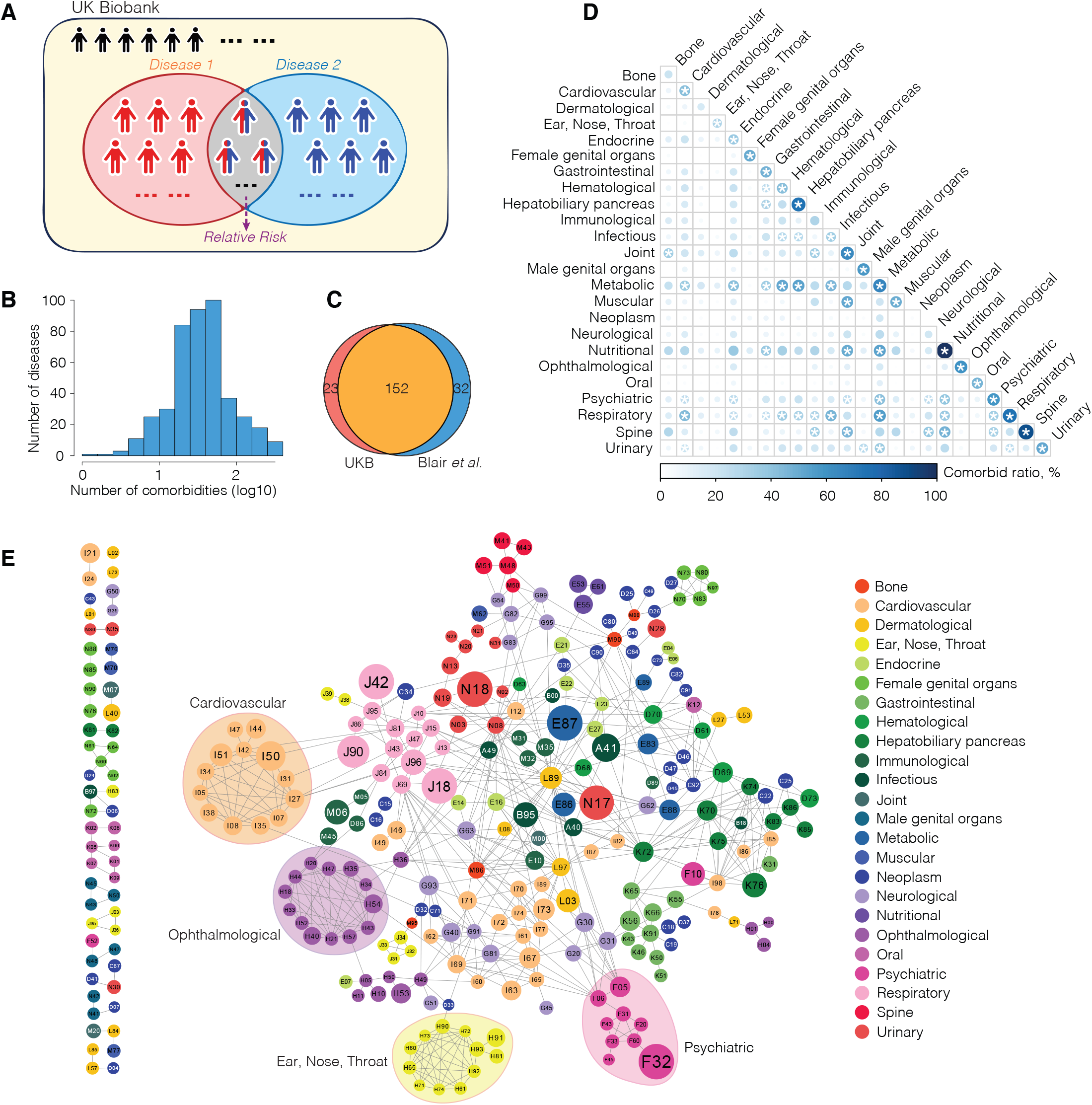
Comorbidities identified in UKB. **a** Schematic illustration of how RR is calculated for each pair of diseases. **b** The distribution of the number of comorbidities for UKB diseases. **c** Comparison of the comorbid relationships identified by us (“UKB”) and by Blair et al. **d** Disease comorbidity tendency intra- and inter-physiological categories. Color and size of the circles represent the proportions of comorbidities in all disease-pairs within a category or between two categories. The deeper the color and the larger the size of a circle, the higher the proportion is. Star represents adjusted *P*-value < 0.05 (FDR corrected). **e** The high confidence comorbidity network constructed by only including comorbidities with RR > 15. Each node represents a disease, and each edge represents a comorbid relationship between two diseases. The color code of a node represents the category of the disease. The size of each node is proportional to the number of its comorbidties (not restricted to RR > 15).

To validate the credibility of the comorbid relationships found through our analysis, we firstly compare them to the comorbid relationships identified in a recent study by Blair *et al* [24]. Blair *et al*. studied the comorbid relationships between Mendelian diseases (14 out of the 439 diseases used in our study) and complex diseases (62 out of the 439 diseases). Among the comorbid relationships they identified, 184 reach the prevalence criteria set for our analysis (for each comorbid relationship, patients with both diseases have to exceed 1% of patients for at least one of the two diseases). In comparison, the number of comorbid relationships we find between the 14 Mendelian diseases and 62 complex diseases is 175, 152 (86.9%) of which are among the comorbid relationships found by Blair *et al*. (Fig. 1c). We examine the 32 comorbid relationships identified by Blair *et al*. but missed by us, and find that 23 of them fulfill RR>1 and are significant in single test (*P*-value < 0.05), but fail to pass the multi-test correction. This is possibly due to that in our analysis, we test more than 12000 pairs of diseases (RR > 1) at once, which incurs a strict multi-test correction with a trade-off between type I error (false positives) and type II error (false negatives). In other words, our analysis may err on the more conservative side. On the other hand, among the 23 comorbid relationships identified by us but not by Blair *et al*., 22 of them have literature evidence supporting their existence, indicating their potential validity (Additional file 2: Table S3). Additionally, we also compare the comorbidities identified by us to those identified by Jensen *et al* [25] (Additional file 3: Supplementary Text). The comparison results show that the comorbidities identified by us and Jensen *et al*. have a significant overlap (*OR*=6.8, *P*=0, Fisher exact test). Taken together, we consider that our results can be confirmed by previous results, and are of high reliability. Importantly, benefiting from the long follow-up time (25 years) and the extensive coverage of diseases provided by UKB hospital inpatient data, the collection of comorbidities reported here is the largest one of common disease comorbidity by far, and thus provide an atlas of comorbidities useful for further analysis.

### Prevalent comorbidity intra- and inter-physiological systems

In the classic Human Disease Network (HDN), diseases are connected if they share at least one disease-associated gene [22]. We have observed several clusters in HDN formed by diseases affecting the same physiological systems, such as cardiovascular diseases and nutritional diseases. This inspires us to explore whether diseases affecting the same physiological system also tend to be comorbidity. We divide the 439 diseases into 24 categories, mostly according to their affected physiological systems while also considering their origins (e.g. “Neoplasm”) (Additional file 2: Table S1), and calculate the disease comorbidity tendency among the 24 categories (see Methods). As expected, we find significantly prevalent comorbidity within 19 out of 24 categories (Fig. 1d). For example, 89% (32/36) of all possible disease-pairs within the “Spine” category are comorbidities (adjusted *P*= 5e-5), and all the 4 nutritional diseases are comorbid with one another (adjusted *P* = 8.9e-3). To get a visual and more confident observation, we construct a high-confidence comorbidity network comprised of comorbidities with RR>15 (Fig. 1e), and are able to observe small and large clusters of comorbidities affecting the same physiological system, many of which are supported by previous studies ⍰ for examples, comorbidity clusters affecting the “Cardiovascular” [39], the “Ophthalmological” [40], the “Ear, Nose, Throat” [41], and the “Psychiatric” [42] categories. These findings suggest shared mechanisms for diseases affecting the same physiological systems.

Interesting, apart from the intra-category comorbidity, we also find significantly more comorbid relationships between 41 pairs of different categories (Fig. 1d). Diseases affecting the “Respiratory” system are significantly more likely to be comorbid with diseases from 11 other categories, followed by diseases affecting the “Metabolic” system with diseases from 10 other categories, both suggesting shared etiologies beyond the boundaries of physiological systems. Moreover, metabolic diseases have overall the highest comorbid rate with diseases of other categories, which is consistent with many reports on the involvement of altered metabolism in a wide range of diseases [43, 44]. It is noteworthy that sometimes the significant cross-category comorbidity patterns are mediated by a small number of diseases that have a large number of comorbid partners. For example, in the high-confidence comorbid network (Fig. 1e), psychiatric disorders comorbid with neurological disorders predominantly through F05 (“Delirium, not induced by alcohol and other psychoactive substances”) and F06 (“Other mental disorders due to brain damage and dysfunction and to physical disease”). In fact, for 33 out of the 41 significant comorbid category-pairs, more than 50% of the inter-category comorbid relationships are mediated through no more than 3 “hub” diseases. Take one of the most centered hubs as an example, E66 (“Obesity”) mediates more than half of the comorbid relationships between the “Nutritional” category and three other categories ⍰ “Psychiatric”, “Spine”, and “Joint”. This is consistent with previous findings that obesity is usually associated with mental, joint, and spinal diseases, such as depressive disorders, anxiety disorders, gout, and spondylosis [45-47]. As a result, understanding the mechanisms underlying comorbidities, especially those mediated by the “hub” diseases, may provide a way forward to understand how they happen, and seek to manage or treat them simultaneously.

### 46% of the comorbidities are genetically interpretable

Previous studies have shown that diseases with more genes or PPIs shared are more likely to be comorbidities [9, 10]. However, it still remains unclear that how many comorbidities share genetic components (deemed as genetically interpretable), and whether there are any specific patterns in shared genetic components for different types of comorbidities. To explore these two questions, we capture the genetic associations of comorbidities at 3 levels, i.e. loci level (SNP and genes), network level (PPI and pathways) and overall genetic architecture level (genetic correlation) (see Methods).

All available GWAS summary statistics based on UKB subjects are collected from geneAtlas [19], covering 332 out of 439 diseases used in this study and comprising 8,212 comorbidies. We find 46% (3,766) of comorbidities have shared genetic components: 147, 1,463, 1,803, and 1,959 comorbidities share SNPs, genes, PPIs, and pathways, respectively; and 1,970 comorbidities have significant genetic correlations (Fig. 2a, Additional file 2: Table S4, S5, S6, S7, S8, see Methods). Comorbidities are significantly more likely to share genetic components, compared with non-comorbidities across all genetic levels as well as their aggregation (Fig. 2a, see Methods). Additionally, we also find that 98%, 70% and 100% of the genetically interpretable comorbidities share significantly more SNPs, genes and pathways than expected, respectively (see Methods). Only 5% of comorbidities share significantly more PPIs than expected, possibly due to the universality of PPIs. Moreover, the genetically interpretable comorbidities have a significant overlap with disease-pairs reported by Park *et al*., which have shared genes, PPIs or co-expressed genes (*P*= 2.4e-6, Fisher exact test), further confirming the genetic associations of comorbidities identified in this study [9]. As the genetic information and the epidemiological information come from the same subjects, we consider our results relatively robust against the usual confounding factors for genetic analysis of comorbidity, such as differences in genetic background. Thus, our results strongly support the existence of genetic predispositions for almost half of the identified comorbid relationships. We have created a queryable online database to facilitate researchers and physicians to explore the comorbidities of interest (https://comorbidity.comp-sysbio.org).

**Fig. 2.**
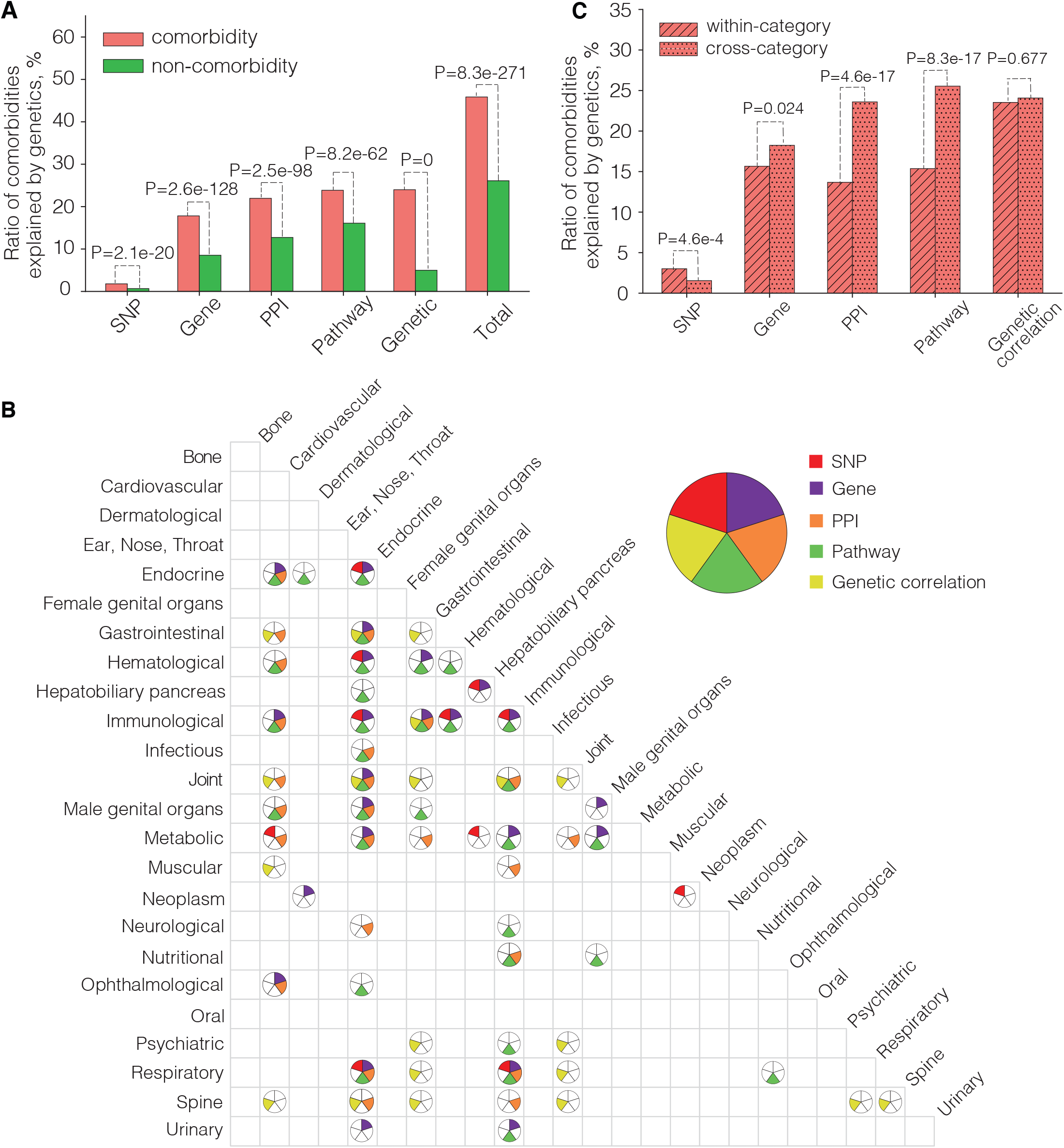
Five types of genetic components shared by comorbidities. **a** Ratios of comorbidities interpretable through SNP, gene, PPI, pathway, and genetic correlation, compared with non-comorbidities. **b** Intra- and inter-category comorbidity that can be significantly interpreted through the five types of genetic components. Each circle is divided into five parts, representing the five types of genetic components. Color-filled parts of each circle represent the types of genetic components that can significantly inteprete the corresponding intra- or inter-category comorbidity. No circle is drawn where none of the five types of genetic components is significant. **c** Ratios of comorbidities of intra- and inter-categories interpretable through SNP, gene, PPI, pathway, and genetic correlation.

We then explore whether there are any specific patterns in shared genetic components for different types of comorbidities, i.e. exploring differences in the types of genetic components mediating intra- and inter-category comorbidity. Overall, we find that comorbidities of 9 (37.5%) intra-categories and 52 (18.8%) inter-categories significantly share genetic components, suggesting a high probability of genetic involvement for comorbidities affecting the same physiological systems (Fig. 2b). Interestingly, as in Figs. 2b and 2c, intra-category comorbidities are slightly more likely to share loci-level genetic components (*P*=4.6e-4 for SNPs; difference not significant for genes after Bonferroni correction; Fisher exact test) compared to inter-category comorbidities, while the latter are more likely to share network-level genetic components (*P*-values =4.6e-17 and 8.3e-17 for PPIs and pathways, respectively). There is no significant difference in how likely comorbidities of intra- and inter-category have genetic correlations. These results suggest that comorbidities affecting the same and different physiological systems may have different biological origins ⍰ with the former tend to directly originate from pleiotropic loci and the latter tend to indirectly originate from converged functions.

The above statistical observation is best illustrated by the diseases affecting male genital organs (Fig. 2b). The comorbidities within “Male genital organs”category tend to share genes (adjusted *P*-value = 4.1e-2, FDR corrected). In fact, 50% (8/16) of the comorbidities within this category share disease-associated genes. 20 genes are involved in these intra-category comorbid relationships, and are mainly related to the human leukocyte antigen (HLA) complex (such as *HLA-DQA1* and *HLA-DRB1*), histone clusters (such as *HIST1H1B* and *HIST1H2AJ*), and tumors (such as *TERT* and *NOTCH4* for prostate cancer) [48, 49]. In contrast, comorbid relationships of inter-category involving the “Male genital organs” category tend to share pathways (72/142, 51%). The KEGG pathway “cell adhesion molecules cams” is shared by half (36/72) of these cross-category comorbid relationships, followed by “antigen processing and presentation”, which is shared by 32 of these comorbidities.

To summarize, based on matched genetic and epidemiological data, we find that almost half (46%) of comorbidities identified in this study are genetically interpretable, indicating a strong genetic role in the origin of comorbidity. Among these genetically interpretable comorbidities, the intra-category and inter-category ones tend to share genetic components in different levels (loci VS. network), suggesting their different biology origins.

### Genetically interpretable comorbidities converge on cell immunity, protein metabolism, and gene silencing

To enhance the understanding about the biological mechanisms of comorbidities, we conduct functional analyses of the loci level and network level genetic components. The overall genetic architecture is not analyzed here, as it reflects statistical correlations but not detailed functions (see Discussion).

We firstly examine the genome-wide distribution and the deleteriousness of the comorbidity-SNPs (see Methods). We find that comorbidity-SNPs tend to be located in noncoding RNA (*P-values* = 1.2e-44 and 4.1e-147) and intergenic regions (*P-values* = 1.7e-54 and 2.9e-48) (Fig. 3a), but with slightly higher CADD scores (*P-values* = 9.8e-129 and 0) than other-disease SNPs (SNPs that are disease-associated but not shared by comorbidities) and non-disease SNPs (Fig. 3b). These results suggest that comorbidity-SNPs are slightly more deleterious, possibly through playing important roles in gene transcriptional regulation [50]. We have also examined the effects of comorbidity-SNPs on splicing by dbscSNC splicing score [35], but found no difference among the comorbidity-SNPs, other disease-SNPs and non-disease SNPs (Additional file 1: Fig. S4). Additionally, we find that 73% of the comorbidity-SNPs locate in a small region of the genome ⍰ the HLA region (chr6:29,691,116–33,054,976), and 51% of the comorbidities which are interpretable through SNPs share at least one HLA-region SNP. The HLA region is well known for its high degree, long-ranged LD blocks, which may help explain the pleiotropy of these SNPs in comorbidities [51]. SNPs in this region have been previously predicted to be relevant for multiple autoimmune diseases through disrupting the regulation of immune-related genes [50]. Consistent with this, most of the top comorbidities with the largest number of shared HLA-region SNPs contain autoimmune diseases or contain a significant autoimmune-related origin. For example, E03 “Other hypothyroidism”, J45 “Asthma”, K90 “Intestinal malabsorption”, E10 “Insulin-dependent diabetes mellitus”, and G35 “Multiple sclerosis”. Finally, we test whether the genomic location and CADD score distributions of comorbidity-SNPs are mainly determined by the HLA-region variants. After removing the HLA-region SNPs, comorbidity-SNPs are still overrepresented in noncoding RNA regions (*P-values*=3.5e-47 and 1.2e-61), and still have significantly higher CADD scores (*P-values*=0.02 and 1.5e-15) than other-disease SNPs and non-disease SNPs, but are no longer overrepresented in intergenic regions (Additional file 1: Figs. S5A, S5B). We conclude that comorbidity-SNPs, no matter whether in the HLA region or not, are slightly more deleteriousness and more likely to locate in noncoding RNA region than other SNPs.

**Fig. 3.**
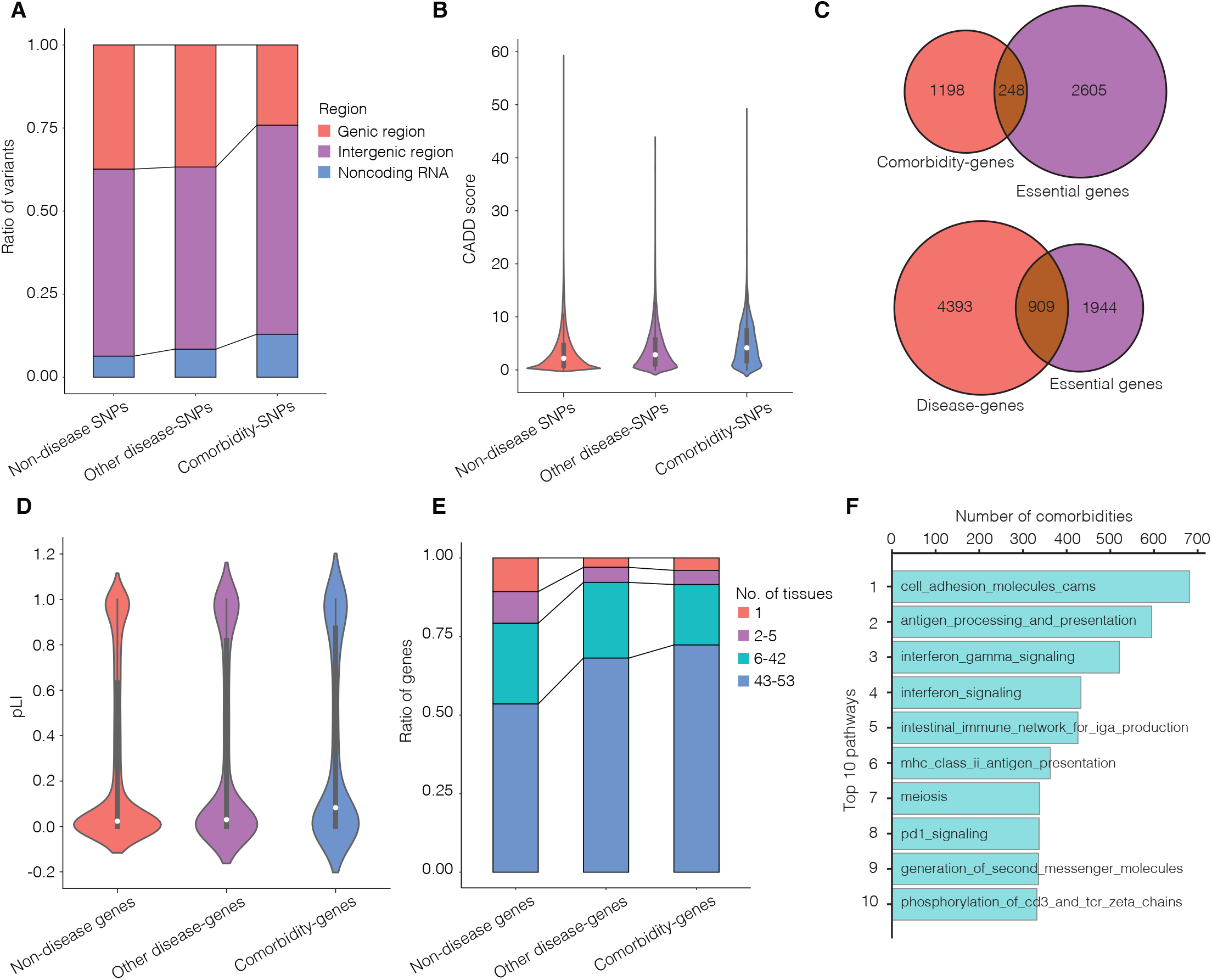
Characteristics of the genetic components shared by comorbidities. **a** The ratios of SNPs located in genic region, intergenic region, and noncoding RNA region for comorbidity-SNPs, other disease-SNPs, and non-disease SNPs. **b** CADD score distributions for comorbidity-SNPs, other disease-SNPs, and non-disease SNPs. **c** Overlaps between comorbidity-genes and essential genes, and between disease-genes and essential genes. **d** The pLI distributions of comorbidity-genes, other disease-genes, and non-disease genes. **e** The ratios of genes expressed in few or many types of tissues, for comorbidity-genes, other disease-genes, and non-disease genes. **f** Top ten pathways that are shared by the largest numbers of comorbidities.

Goh *et al*. reported that most (78%) disease-genes (i.e. genes associated with at least one disease recorded in Online Mendelian Inheritance in Man) are not essential genes critical for survival, and that disease-genes are less likely to be housekeeping genes that express in all tissues [22]. We then test whether our comorbidity-genes behave similarly or differently. Here, we obtain 2,852 essential genes, which are human orthologs of mouse genes whose disruptions are embryonically or postnatally lethal (see Methods). We find that only 17% of the disease-genes (identified by us) and 17% of the comorbidity-genes are essential (Fig. 3c), although disease-genes and comorbidity-genes are more enriched in essential genes than non-disease genes (*P-values* = 2.9e-38 and 8.8e-15, respectively). Essential genes are reported to have a tendency to encode hub proteins in human interactome, and play an important role in maintaining normal developmental and/or physiological function [22]. These results indicate that most comorbidity-genes are functionally peripheral in human interactome, and their mutations are compatible with survival into reproductive years so that these comorbidity phenotypes are conveyed in a population. Although most comorbidity-genes are not essential, we find a higher probability of loss of function intolerance (pLI) for comorbidity-genes, compared to other disease-genes as well as non-disease genes (*P-values* = 0.01 and 3.6e-11, respectively, *t* test; Fig. 3d). Removing the essential genes, this trend remains unchanged, suggesting the higher pLI distribution of comorbidity-genes is not just due to the essential genes (Additional file 1: Fig. S6). To examine whether comorbidity-genes tend to be housekeeping genes, we summarize the number of tissues each gene is expressed in based on the gene expression data for 53 tissues in GTEx [37]. We find that comorbidity-genes are more likely to be expressed in the majority of tissues, compared to other disease-genes and non-disease genes (*P* = 4.7e-4 and 8.1e-44, respectively, two-sided Mann–Whitney U-test; Fig. 3e). Considering the high pleiotropy of HLA regions, we recalculate the properties of the comorbidity-genes after removing HLA variants. In this case, we find that most comorbidity-genes are still nonessential and comorbidity-genes still tend to have higher pLI (*P-values*=0.04 and 2.9e-10) and express in more tissues(*P-values*=4.4e-3 and 1.2e-32) compared with other-disease genes and non-disease genes (Additional file 1: Figs. S5C, S5D, S5E). As a result, we consider that most comorbidity-genes are very important for normal biological mechanisms, though most of them are not essential for survival, and disrupted comorbidity-genes may have clinical consequences affecting slightly more tissues than other disease-genes.

For the network level genetic components, the genes involved in the top 10 PPIs shared by the most numbers of comorbidities are significantly enriched in GO terms related to biological processes of gene silencing and protein metabolism including localization, acetylation, ubiquitination, catabolism (see Methods). These 10 PPIs account for 18% of the PPI interpretable comorbidities. Moreover, as shown in Fig. 3f, the top 10 pathways shared by the most numbers of comorbidities are predominantly immune-related processes, and correspond to 56% of all the pathway interpretable comorbidities. Most of the top pathway involved diseases are autoimmune or inflammatory diseases, such as J45 “Asthma”, K20 “Oesophagitis”, M06 “Other rheumatoid arthritis”, L40 “Psoriasis”, and E10 “Insulin-dependent diabetes mellitus”. The findings based on network level genetic components suggest a phenomenon that a significant portion of genetically interpretable comorbidities may converge on a handful of biological mechanisms, with the most common mechanisms related to cell immunity, gene silencing and protein metabolism. This phenomenon is further supported by the loci level genetic components: the top 10 genes can interpret as much as 41% of the gene interpretable comorbidities, and are enriched in immune-related GOs, such as “interferon gamma mediated signaling pathway”, “antigen processing and presentation of peptide antigen”, and “regulation of T cell mediated cytotoxicity”. Moreover, after removing HLA-region SNPs, we still find a few genetic components interpreting many comorbidities (the top 10 SNPs, genes, PPIs and pathways can interpret 23%, 30%, 15%, 34% of comorbidities interpretable through SNP, gene, PPI, pathway, respectively.). The top enriched GO terms are “protein localization to chromosome telomeric region” and “beta-catenin-tcf complex assembly”, and the top enriched pathways are “rna pol i rna pol iii and mitochondrial transcription” and “meiosis” (Additional file 1: Figs. S5F).

### “Hub” disease-mediated genetically interpretable comorbidity modules

The fact that a small number of genetic components can interpret a large portion of the genetically interpretable comorbidities, inspires us to examine whether the “small world” phenomenon exist in the shared genetic architectures of those comorbidities. Therefore, we construct two comorbidity networks by connecting comorbid diseases that share loci level genetic components (denote as the LG-network) and network level genetic components (denote as the NG-network), respectively. As expected, for the LG- and NG-networks, their degrees follow the power-law distribution (Additional file 1: Fig. S7), and they have the attributes of small worldness based on their average clustering coefficient and average shortest path length (*sigma*=1.15 and 1.03, respectively; see Methods). In previous sections, we have shown that many of the inter-category comorbidities are mediated through hub diseases. Given the “small world” properties of the LG- and NG-networks, we hypothesize that there are comorbidity modules in the two networks, possibly featured by hub diseases and specific genetic components. In order to test this hypothesis, we perform network decomposition to detect comorbidity modules by the Louvain algorithm [38].

We first arbitrarily define nodes (diseases) connected with more than 25% of all nodes in each network as “universal hub diseases”. It is not appropriate to assign the “universal hub diseases” into any single comorbidity module, as each module usually contains far fewer than 25% of all nodes. Seven and thirteen “universal hub diseases” are found for the LG-network and the NG-network, respectively (Fig. 4, Additional file 2: Table S9). The “universal hub diseases” are usually known to co-occur with many diseases. For example, Hypertension (I10) connect to 235 (80%) diseases in the NG-network, and is well-known for having heavy comorbidity burdens [52].

**Fig. 4.**
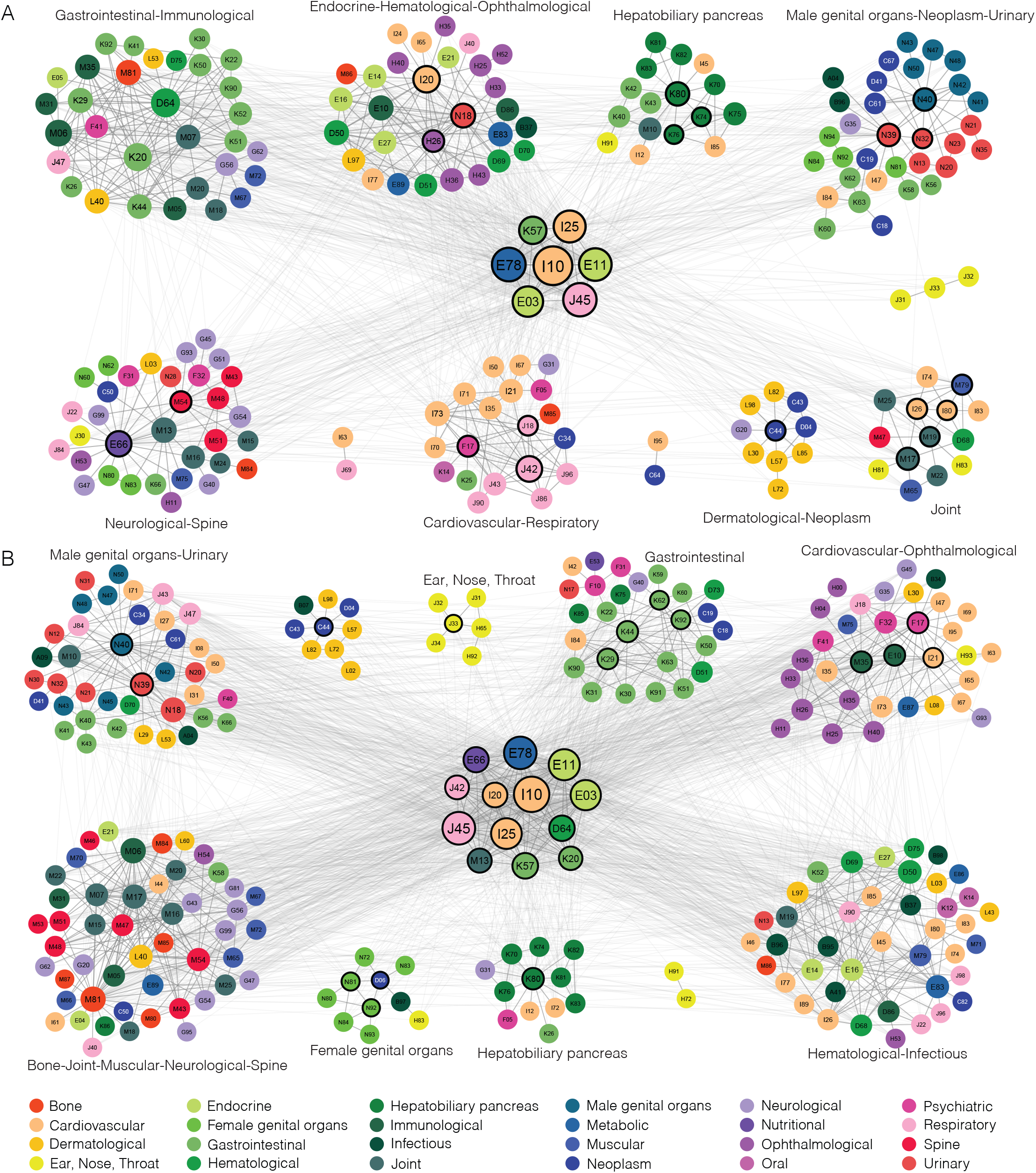
Loci-level (a) and Network-level (b) genetically interpretable comorbidity networks. **a, b** Each circle represents a disease. Colors of the circles (diseases) correspond to the categories of the diseases. “Universal hub diseases” are located at the center of each network, and diseases that belong to the same comorbidity module are grouped close to each other. For each module, the featured categories are annotated. The black border of the nodes indicates that they are the local hub diseases of the comorbidity modules. Diseases that belong to neither universal hub diseases nor a comorbidity module are not included in this figure.

Next, based on the remained nodes that are not “universal hub diseases”, we have detected 11 comorbidity modules for the LG-network and 10 comorbidity modules for the NG-network (Modularity *Q* = 0.40 and 0.32, respectively) (Figs. 4a and 4b, Additional file 2: Table S10, S11). Overall, the module sizes (number of nodes in a module) range from 2 to 49, and the LG- and NG-networks have 8 and 9 comorbidity modules whose sizes are larger than 5, respectively. Each comorbidity module is assigned with “featured categories”, which are the categories that the diseases in this module are significantly overrepresent (Fisher exact test, adjusted *P*-value <= 0.05, FDR corrected). Within each module (size > 5), if diseases with top 3 largest number of within-module comorbidities mediate more than half of the within-module edges, we define them as “local hub diseases” of the module. As shown in Table 1, we have identified “featured categories” for all comorbidity modules in the LG- and NG-networks, and “local hub diseases” for 7 and 7 comorbidity modules in the LG- and NG-network, respectively. We find that most local hub diseases belong to the featured categories of their modules, highlighting the prevalence of genetically interpretable comorbidities within the same physiological system. Nonetheless, most modules have more than one featured category, demonstrating that genetically interpretable comorbidities are not limited by physiological boundaries.

**Table 1.**
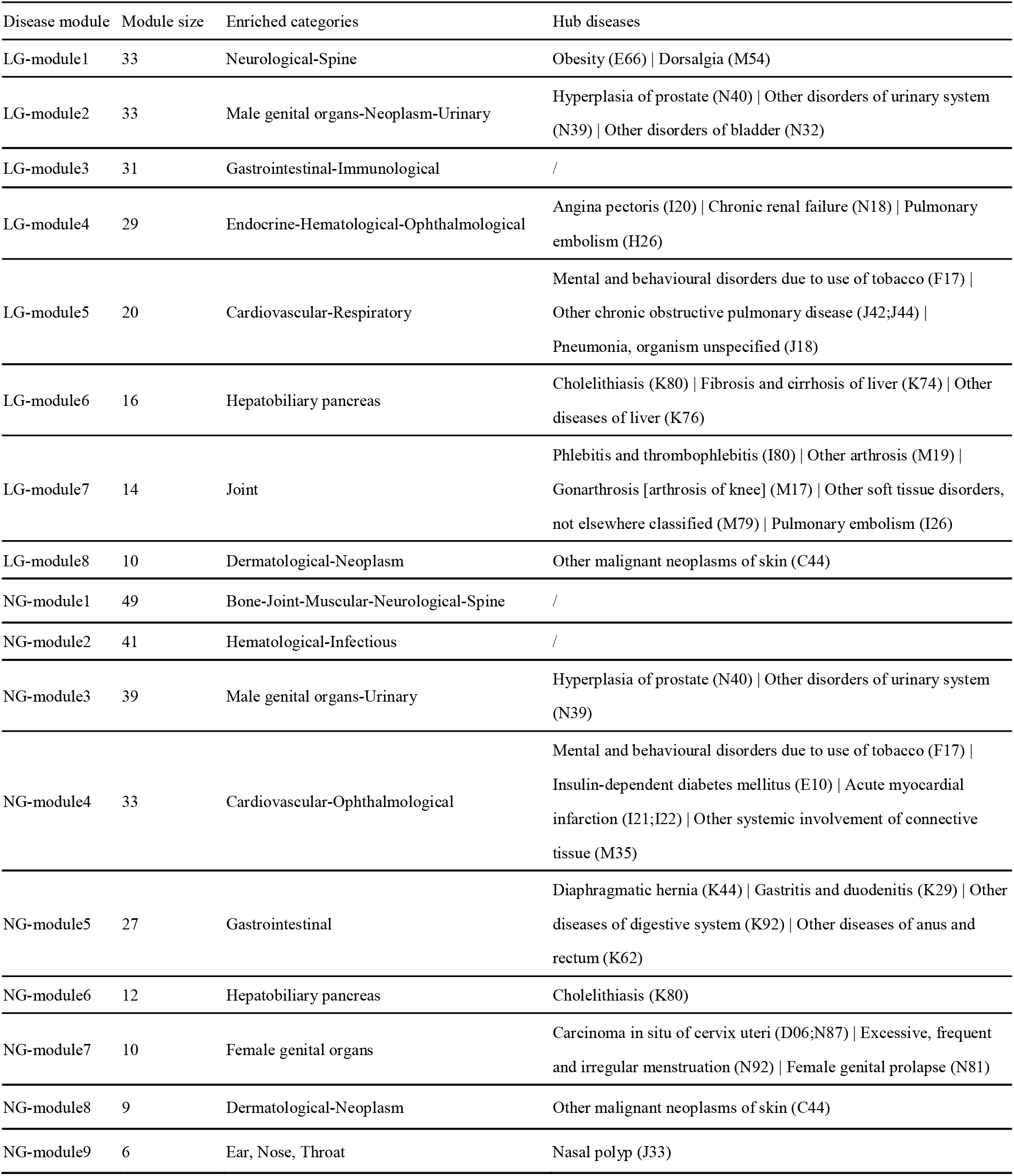
Genetically interpretable comorbidity modules and their hub diseases.

We will describe several cases to illustrate how the network and module structures can help us understand the genetics underlying large numbers of comorbid relationships. Firstly, some categories are consistently grouped together. In both networks, we identify modules that feature Male genital organs-Urinary, Dermatological-Neoplasm, and Neurological-Spine categories, supporting the existence of genetic associations between the involved comorbidities from multiple genetic levels (Table 1). Secondly, modules can help distinguish different comorbidity tendencies and the corresponding genetic mechanisms among diseases of the same categories. In the LG-network, Neoplasm diseases are featured in two modules ⍰ LG-module2 (Male genital organs-Neoplasm-Urinary) and LG-module8 (Dermatological-Neoplasm). Neoplasm diseases in LG-module2 are mainly prostate (C61), bladder (C67), urinary organs (D41) and intestinal (C18,C19;C20) cancers, while Neoplasm diseases in LG-module8 are all skin cancers (C43, C44, D04) (Fig. 5a, Additional file 2: Table S10). Except for *TERT* and *CLPTM1L*, genes shared by Neoplasm diseases and other diseases in the two modules are different, reflecting diverse mechanisms among Neoplasms related to different tissues. Thirdly, hub diseases may provide a new perspective for understanding comorbidity with different categories of diseases. For example, as shown in Fig. 5b, the Psychiatric disorder F17 “Mental and behavioral disorders due to use of tobacco” is a hub diseases of LG-module5 (Cardiovascular-Respiratory), and is comorbid with 5 respiratory diseases (emphysema (J43), pyothorax (J86;J93), respiratory failure (J96), chronic obstructive pulmonary disease (COPD, J42;J44), pneumonia (J18)), 3 cardiovascular diseases (atherosclerosis (I70), aortic aneurysm and dissection (I71), peripheral vascular diseases (I73)), and lung cancer (C34). The most often genes shared by these comorbidities are *IREB2* and *CHRNA3*, located in 15q25, a well-known region for association with COPD, lung cancer, and smoking [53]. The *IREB2* gene encodes an iron-responsive element-binding protein (IRP) that regulates the iron metabolism [53]. *CHRNA3* encodes the neuronal nicotinic acetylcholine receptor, and its mutation is associated with lung function and COPD severity in ever-smokers [54]. Though there are many other possible pathways individually associated with the above diseases, our analysis indicates that iron metabolism and the neuronal nicotinic acetylcholine receptor pathway may be the top candidates to examine when study the comorbidity of these diseases with F17. Lastly, “universal hub diseases” connect to multiple modules, sometimes through different genetic components. In the NG-network, the universal hub disease Obesity (E66) have multiple connections to NG-module1 (Bone-Joint-Muscular-Neurological-Spine) and NG-module6 (Hepatobiliary pancreas) (Fig. 4b). Pathways shared by Obesity and Hepatobiliary pancreas diseases are mostly related to lipoprotein metabolism, such as “lipid digestion mobilization and transport”, “fatty acid triacylglycerol ketone body metabolism”, “cytosolic sulfonation of small molecules”, “mitochondrial protein import”, while also related to biological oxidations, myogenesis etc (Fig. 5c, Additional file 2: Table S11). In comparison, pathways shared by Obesity and Bone, Joint, Muscular, Neurological, Spine diseases from NG-module1 are mostly related to immunity, cell adhesion and transcription. In summary, the genetically interpretable network and modules can provide insights through hub diseases to the understanding of the molecular mechanisms underlying comorbidity, and may help prioritize target genes and pathways for designing new treatment.

**Fig. 5.**
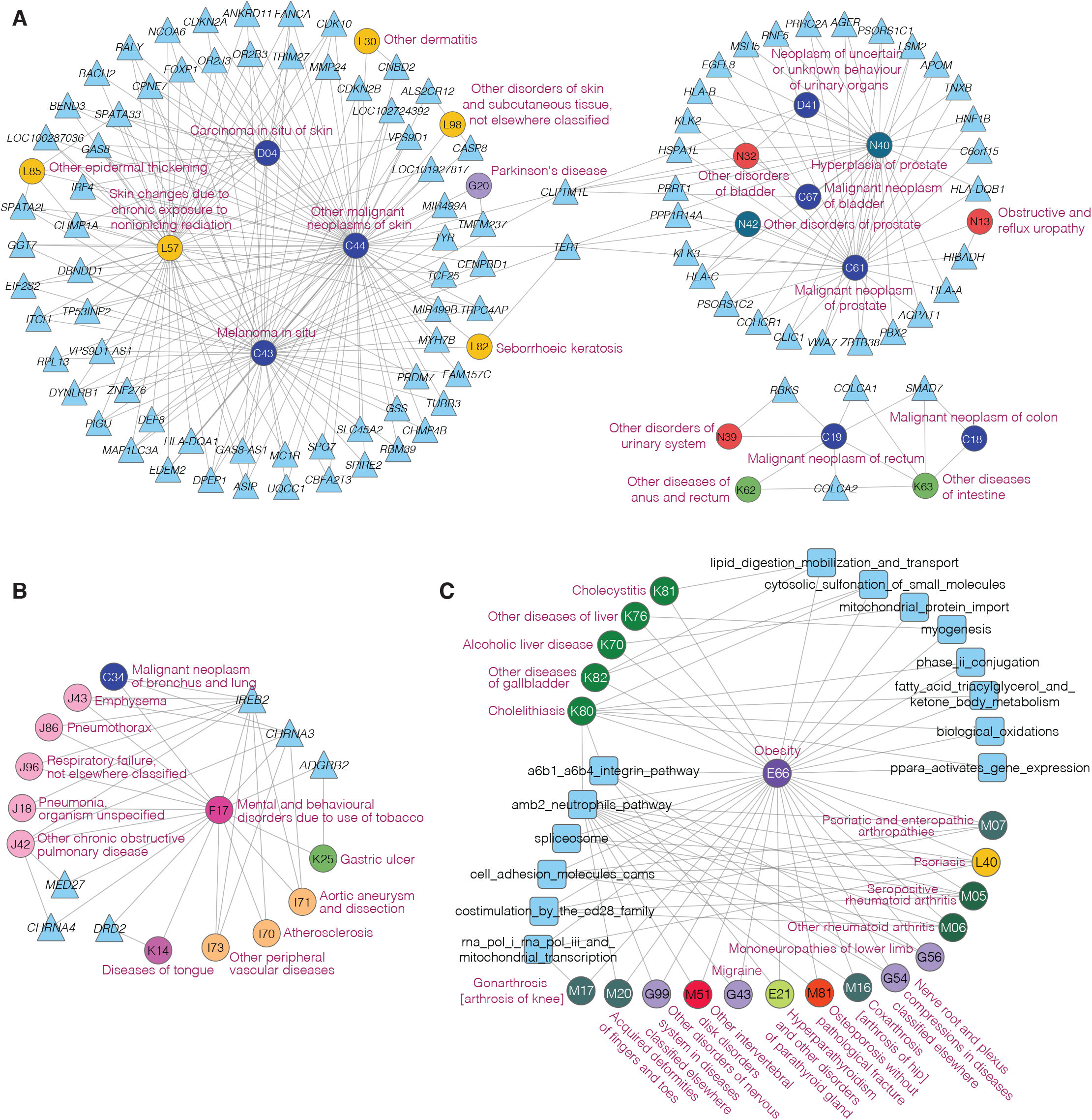
Case studies for genetically interpretable comorbidity networks. **a-c** Circles, triangles, and squares represent diseases, genes, and pathways, respectively. Colors of the circles (diseases) correspond to the categories of the diseases, following the same color code as in Fig. 1. **a** Neoplasm diseases, their comorbid diseases, and shared genes in LG-module2 and LG-module8. **b** Comorbidities of the hub disease F17 (Mental and behavioural disorders due to use of tobacco) in LG-module5, and their shared genes. **c** Pathways shared by the universal hub disease E66 (Obesity) and its comorbidities in NG-module1 and NG-module6. Only the top 5 pathways shared by the largest numbers of comorbidities are shown.

## Discussion

In this study, we have profiled the comorbid relationships among the common diseases in UKB, and systematically investigated the genetic risks shared by comorbidities. We report an atlas of 11,285 comorbid disease-pairs among 438 common diseases, which is by now the largest in scale. We find that 46% of the comorbidities with available genetic information share genetic components in at least one of the three levels ⍰ loci, network, or overall genetic architecture, and show that comorbidities affecting the same and different physiological systems tend to share different levels of genetic components. Functional analyses show that the loci level genetic components shared by comorbidities tend to be more deleterious (for SNPs) and affect more tissues (for genes), and both loci and network level genetic components mainly converge on cell immunity, protein metabolism, and gene silencing related functions. We also construct two comorbidity networks by genetically interpretable comorbidities, and show that hub diseases mediating most connections among comorbidity modules can provide useful insights into the genetic contributors for comorbidities affecting different physiological systems. Therefore, our results provided a detailed comorbid and genetic landscapes of many common diseases, which may be valuable for guiding the early diagnosis, management, and treatment of comorbidity.

Our results highlight shared genetic predispositions or mechanisms underlying comorbidity, which may provide useful information for drug discovery. Theoretically, it is plausible to repurpose existing drugs that target shared genetic components of a pair of comorbid diseases, to treat the comorbidity of the two diseases. In an exploratory test, we are able to identify 8,458 drug-comorbidity relationships with the drug known to target the comorbidity-genes (Additional file 2: Table S12). Interestingly, some of these drugs are indeed used in the population with the corresponding comorbidity. For example, the gene *EDNRA*, a known target of aspirin, is shared by I20 “Angina pectoris” and I25 “Chronic ischaemic heart disease”, and we find that 65% of the people suffering from both diseases report usage of aspirin in UKB. Moreover, the indication of aspirin for I20 “Angina pectoris” and I25 “Chronic ischaemic heart disease” have been reported by the Comparative Toxicogenomics Database (CTD, http://ctdbase.org/; Supplementary Data 12) [55]. Besides this encouraging case, we also found a surprising case concerning Lansoprazole, which targets *MAPT*, a gene shared by E66 “Obesity” and J84 “Other interstitial pulmonary diseases”. 16.7% of the people suffering from both diseases ever used lansoprazole according to the UKB. We find the indications of lansoprazole only include E66 “Obesity” in CTD, but lansoprazole was reported to be able to induce interstitial lung disease [56], suggesting that some of the comorbidity of E66 “Obesity” and J84 “Other interstitial pulmonary diseases” may be due to the use of lansoprazole. Though very preliminary, these initial results shed light on the possibility that our resource of comorbidities and their shared genetic components may help with drug discovery as well as avoid severe side-effects for treating comorbidity in the future.

Notably, as much as 24% of the 8,218 comorbidities have significant genetic correlations, supporting the polygenic architecture of complex diseases, while nearly half of them can not be readily interpretable at either the loci or the network level (Additional file 1: Fig. S8). This indicates unidentified genetic information within the genetic architecture that may require further investigation, such as the copy number variants (CNVs). CNVs are the structural chromosomal variants greater than 1 kb in size, and usually have dosage effects for genes. Several common diseases have been reported to be associated with rare CNVs, such as Autism, schizophrenia [57, 58]. In addition, we find that genetic correlation is positively and significantly correlated with RR (r = 0.39, *P*-value = 1.9e-72, Additional file 1: Fig. S9A), and with phenotype similarity of comorbid diseases (r = 0.22, *P*-value = 0.03, Additional file 1: Fig. S9B, phenotype similarity pre-calculated by van Driel *et al* [59].). Moreover, phenotype similarity and RR have a positive and significant correlation (r = 0.27, *P*-value = 1.5e-8, Additional file 1: Fig. S9C). These results suggest that genetic architecture might interpret comorbidity by contributing to symptom similarities.

One possible limitation with our study is the sample size. Though the overall sample size is not small, we do not have that many cases for each disease or each comorbid disease-pair. As such, we may fail to identify some comorbid disease-pairs that are disproportionally represented in our dataset. Also, the GWAS analyses might miss variants with very small effects. Nevertheless, our study is the first and largest study that combines the epidemiological and genetic information of the same subjects to explore the genetic components underlying comorbidity. The matched phenotype and genotype data makes our results less affected by population-related confounding factors. Based on our current findings, one interesting future direction is to integrate more samples from other studies, and incorporate more types of data, such as imaging data and quantitative traits, in order to analyze the endophenotypes that bridge diseases and deepen our understanding of the mechanisms of comorbidity.

## Conclusions

In summary, we have performed, for the first time, a systematic analysis of comorbid relations among common diseases as well as their genetic associations based on the matched epidemiological and genetic data of the same subjects. Our results illustrate the comorbidity tendency and the genetic association patterns of comorbidities of intra- and inter-physiological systems, and indicate that the hub diseases and converged biological molecules and functions may be the key for treating comorbidity. An online database of UKB-Comorbidity is created to facilitate researchers and physicians to search the comorbidities of interest.

## Data Availability

The epidemiological data used in the present study is available from UK Biobank with restrictions applied. Data were used under license and thus not publicly available. Access to the UK Biobank data can be requested through a standard protocol (https://www.ukbiobank.ac.uk/register-apply/). GWAS summary statistics used in this study can be accessed from http://geneatlas.roslin.ed.ac.uk. GRCh37.p13 is downloaded from https://www.ncbi.nlm.nih.gov/assembly/GCF_000001405.25/. The CADD scores are downloaded from https://cadd.gs.washington.edu. The dbscSNC scores are downloaded from ftp://dbnsfp:dbnsfp@dbnsfp.softgenetics.com. The GTEx eQTL data are downloaded from https://gtexportal.org/home/datasets. The MAGMA tool can be found at https://ctg.cncr.nl/software/magma. The Mouse Genome Informatics database is at http://www.informatics.jax.org. The MSigDB database is at https://www.gsea-msigdb.org/gsea/msigdb/index.jsp. The OMIM database is at https://omim.org/help/about. The UMLS description can be found at https://www.nlm.nih.gov/research/umls/index.html. The ExAC database is at https://gnomad.broadinstitute.org.

## Acknowledgements

Not applicable.

## Availability of data and materials

The epidemiological data used in the present study is available from UK Biobank with restrictions applied. Data were used under license and thus not publicly available. Access to the UK Biobank data can be requested through a standard protocol (https://www.ukbiobank.ac.uk/register-apply/). GWAS summary statistics used in this study can be accessed from http://geneatlas.roslin.ed.ac.uk. GRCh37.p13 is downloaded from https://www.ncbi.nlm.nih.gov/assembly/GCF_000001405.25/. The CADD scores are downloaded from https://cadd.gs.washington.edu. The dbscSNC scores are downloaded from ftp://dbnsfp:dbnsfp@dbnsfp.softgenetics.com. The GTEx eQTL data are downloaded from https://gtexportal.org/home/datasets. The MAGMA tool can be found at https://ctg.cncr.nl/software/magma. The Mouse Genome Informatics database is at http://www.informatics.jax.org. The MSigDB database is at https://www.gsea-msigdb.org/gsea/msigdb/index.jsp. The UMLS description can be found at https://www.nlm.nih.gov/research/umls/index.html. The ExAC database is at https://gnomad.broadinstitute.org.

## Ethics approval and consent to participate

This work used data from UK Biobank (project 1954).

## Consent for publication

Not applicable.

## Author’s contributions

Xing-Ming Zhao conceived the project. Guiying Dong collected, analyzed the data, and wrote the manuscript. Jingqi Chen and Xing-Ming Zhao supervised the data analyses, and revised the manuscript. Jianfeng Feng provided the UK Biobank data. Jianfeng Feng and Fengzhu Sun give suggestions to the manuscripts.

## Author details

^1^Institute of Science and Technology for Brain-inspired Intelligence, Fudan University, Shanghai, China. ^2^Key Laboratory of Computational Neuroscience and Brain-Inspired Intelligence (Fudan University), Ministry of Education, Shanghai, China. ^3^Molecular and Computational Biology Program, University of Southern California, Los Angeles, CA, USA.

## Code availability

All code used for data preparation and analysis are available at https://github.com/Arya-bioinformatics/ukb-comorbidity.

## Supplementary information

**Additional file 1: Supplementary Figures and Tables**. Number of comorbidities for diagnosis time windows of no, same day, <half year, < 1 year, < 2 years, <3 years, <4 years, <5 years **(Fig. S1)**. Size distribution of the Reactome pathways, KEGG pathways, BioCarta pathways and PID pathways (top). Disease and comorbidity pathway size distribution of Reactome and non-Reactome when only using pathways with size <= 200 (bottom) **(Fig. S2)**. Correlation between prevalences and number of comorbidities **(Fig. S3)**. Splicing score of comorbidity-SNPs, other disease-SNPs and non-disease SNPs **(Fig. S4)**. Characteristics of the genetic components shared by comorbidities when removing HLA-region variants **(Fig. S5)**. pLIs of comorbidity-genes, other disease-genes and non-disease genes when removing essential genes **(Fig. S6)**. Degree distributions of diseases in comorbidity networks that share loci and network level genetic components **(Fig. S7)**. Comorbidity overlap interpreted by SNPs, genes, PPIs, pathways and genetic correlations **(Fig. S8)**. Correlations among genetic correlation (rg), relative risk (RR), and phenotype similarity (PheSim) of comorbidities. **(Fig. S9)**.

**Additional file 2: Supplementary Tables**. Common disease summary information **(Table S1)**. Comorbidity among common diseases in UK Biobank **(Table S2)**. UK Biobank comorbidity supported by other studies but not Blair’s results **(Table S3)**. Comorbidities interpreted by SNPs **(Table S4)**. Comorbidities interpreted by genes **(Table S5)**. Comorbidities interpreted by PPIs **(Table S6)**. Comorbidities interpreted by pathways **(Table S7)**. Comorbidities interpreted by genetic correlations **(Table S8)**. Summary of universal hub diseases in LG- and NG-networks **(Table S9)**. Comorbidity modules in LG-network **(Table S10)**. Comorbidity modules in NG-network **(Table S11)**. Comorbidity-drug-prescription relations derived from comorbidity genes, drug targets and UK Biobank self-report medications **(Table S12)**.

**Additional file 3: Supplementary Text**.

