## Additional files for "A global overview of genetically interpretable comorbidities among common diseases in UK Biobank": Additional file 1.docx

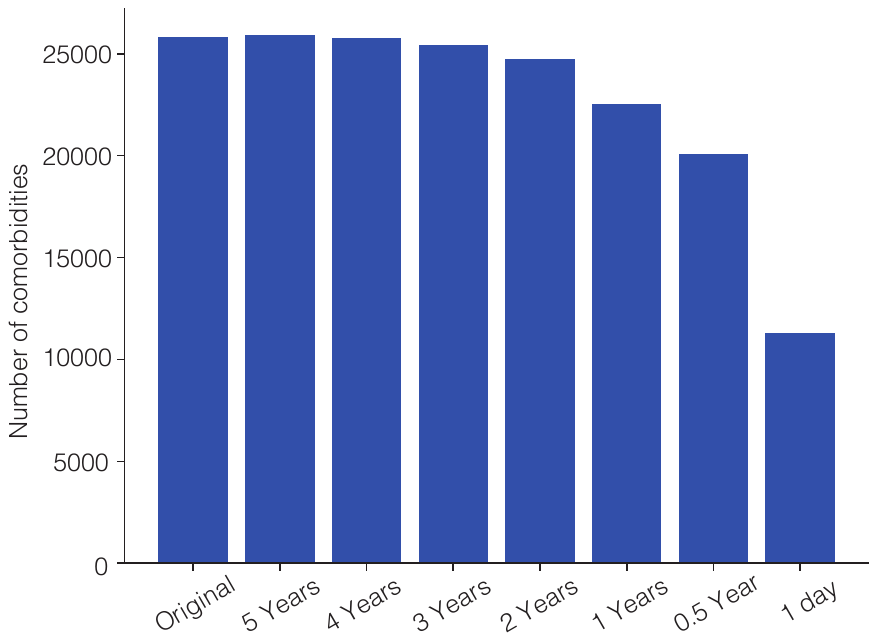

**Fig. S1. Number of comorbidities for diagnosis time windows of no, same day, <half year, < 1 year, < 2 years, <3 years, <4 years, <5 years.**

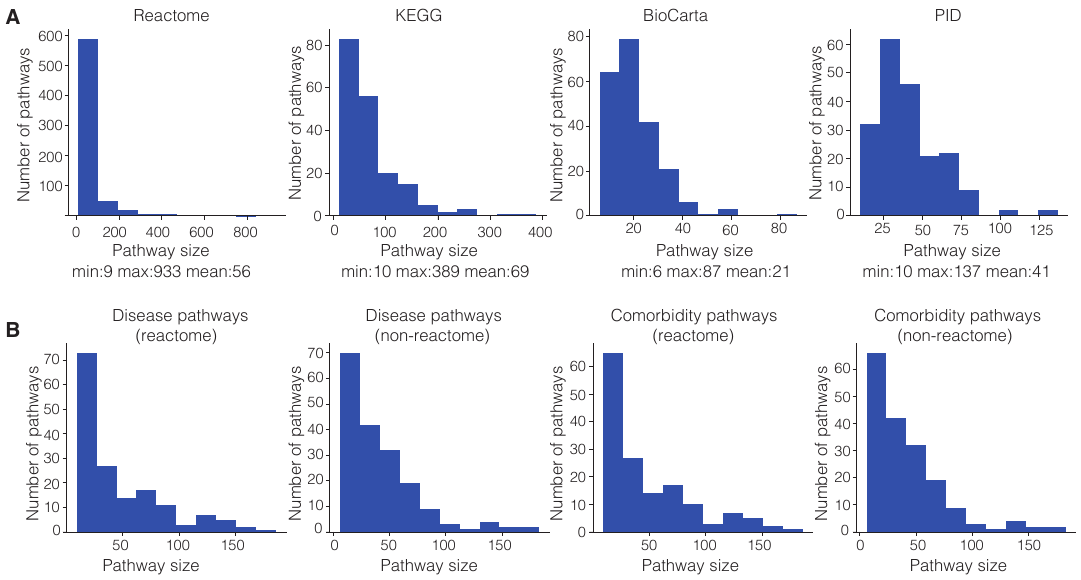

**Fig. S2. Size distributions of the pathways. A** Size distribution of Reactome pathways, KEGG pathways, BioCarta pathways and PID pathways. **B** Size distribution of disease and comorbidity pathways (reactome and non-reactome) bu using pathways with size <= 200 for enrichment analysis.

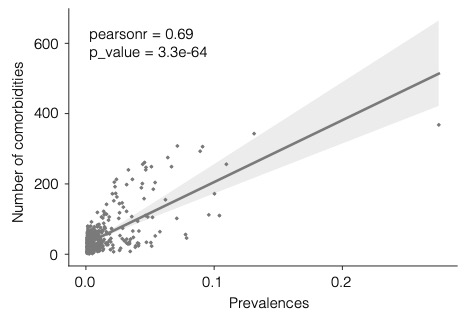

**Fig. S3. Correlation between prevalence and number of comorbidities.**

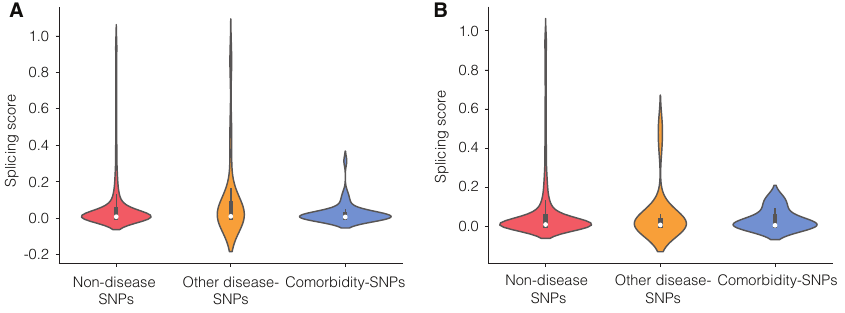

**Fig. S4. Splicing scores of SNPs.** **A** Splicing scores of non-disease SNPs, other disease-SNPs and comorbidity-SNPs. **B** Splicing scores of non-disease SNPs, other disease-SNPs and comorbidity-SNPs after removing HLA-region SNPs.

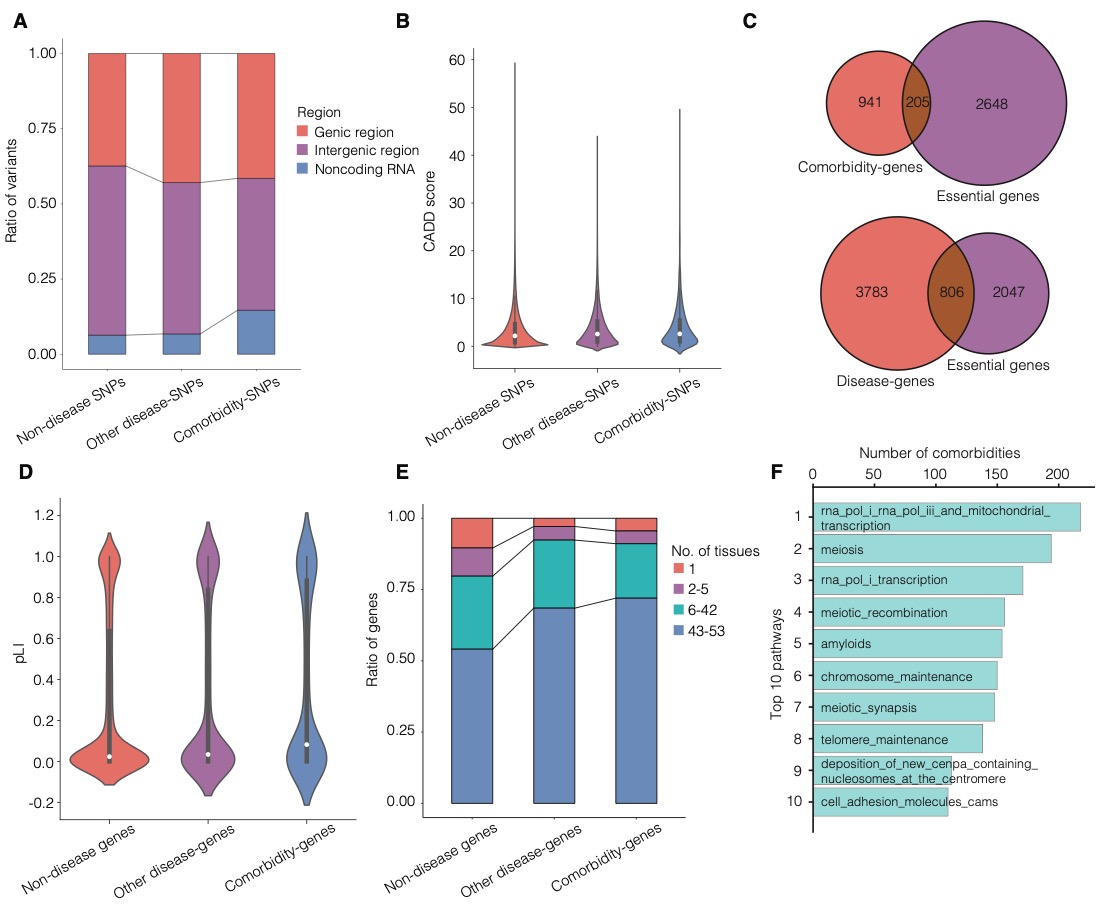

**Fig. S5. Characteristics of the genetic components shared by comorbidities after removing HLA-region SNPs. A** The ratios of SNPs located in genic, intergenic, and noncoding RNA regions for comorbidity-SNPs, other disease-SNPs, and non-disease SNPs. **B** CADD score distributions for comorbidity-SNPs, other disease-SNPs, and non-disease SNPs. **C** Overlaps between comorbidity-genes and essential genes, and between disease-genes and essential genes. **D** The pLI distributions of comorbidity-genes, other disease-genes, and non-disease genes. E The ratios of genes expressed in few or many types of tissues, for comorbidity-genes, other disease-genes, and non-disease genes. F Top ten pathways that are shared by the largest numbers of comorbidities.

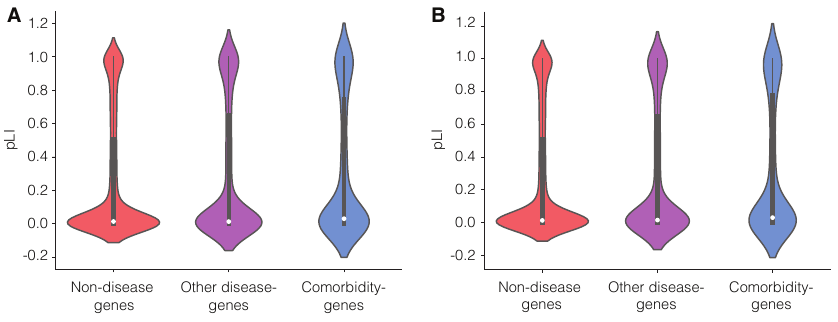

**Fig. S6. pLI scores of genes. A** pLI scores of non-disease genes, other disease-genes and comorbidity-genes. **B** pLI scores of non-disease genes, other disease-genes and comorbidity-genes after removing HLA-region SNPs.

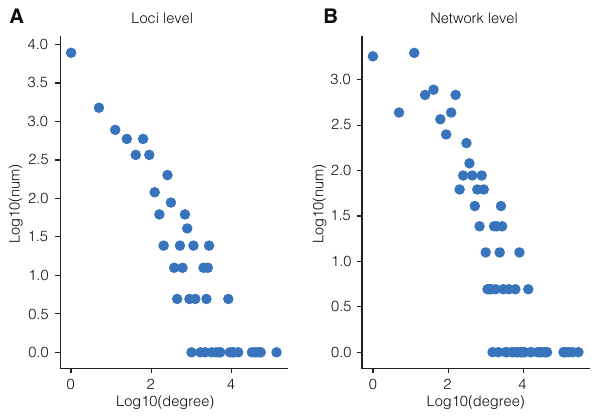

**Fig. S7. Node (disease) degree distributions of LG-network (A) and NG-network (B).**

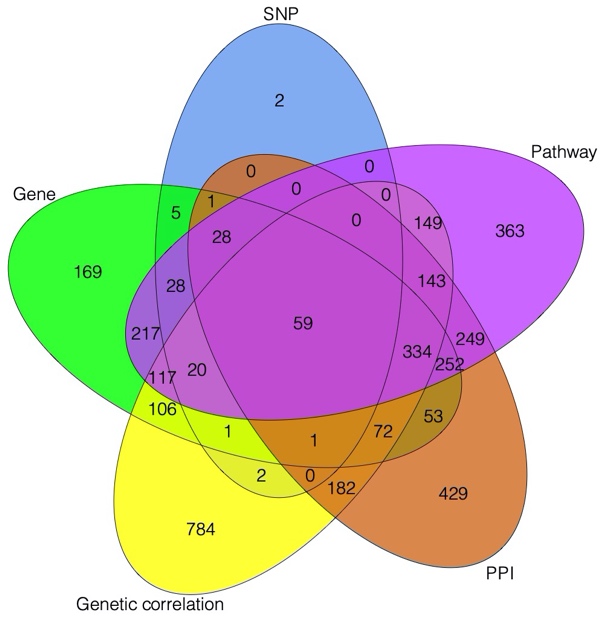

**Fig. S8.** **Comorbidity overlap interpreted by SNP, gene, PPI, pathway and genetic correlation.**

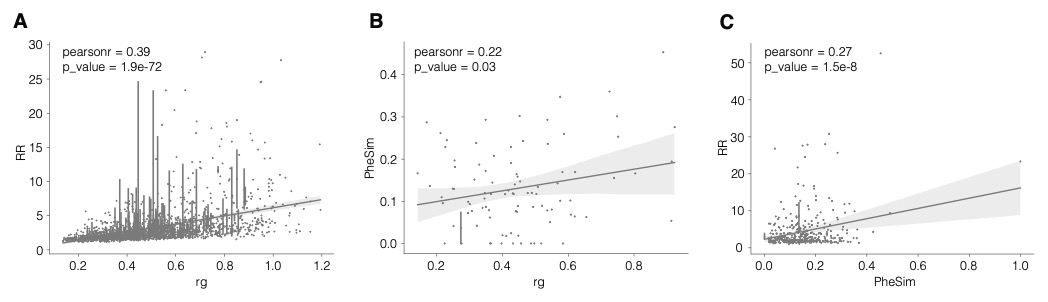

**Fig. S9. Correlations among genetic correlation (rg), relative risk (RR), and phenotype similarity (PheSim) of comorbidities. A** Pearson correlation coefficient calculated between genetic correlation (rg, x-axis) and relative risk (RR, y-axis). 6 dots with RR > 30 are removed for clarity of the figure. **B** Pearson correlation coefficient calculated between genetic correlation (rg, x-axis) and phenotype similarity (PheSim, y-axis). Phenotype similarity was pre-calculated by van Driel et al. (van Driel et al., 2006). 1 dot with PheSim > 0.6 is removed for clarity of the figure. **C** Pearson correlation coefficient calculated between phenotype similarity (PheSim, x-axis) and relative risk (RR, y-axis). 1 dot with RR > 60 is removed for clarity of the figure.

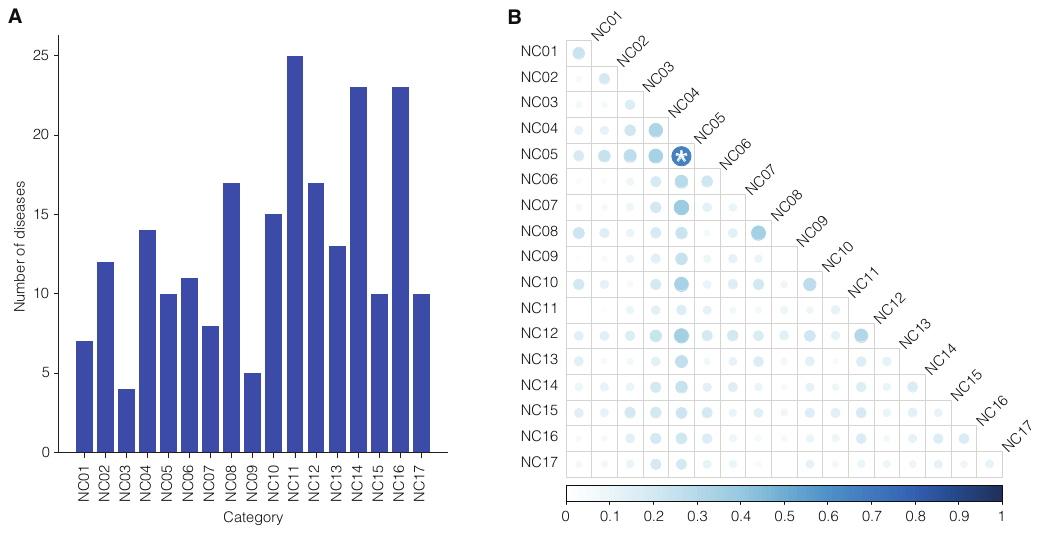

**Fig. S10.** Disease comorbidity tendency intra- and inter-categories based on new chapters (NC) of diseases given by Zhou *et al***. A The number of ICd10 diseases in each new chapter. B** Disease comorbidity tendency intra- and inter-categories. Color and size of the circles represent the proportions of comorbid relationships in all disease-pairs within a category or between two categories. The deeper the color and the larger the size of a circle, the higher the proportion is. Star represents adjusted P-value < 0.05 (FDR corrected).
