## Additional files for "A global overview of genetically interpretable comorbidities among common diseases in UK Biobank": Additional file 3.docx

***Supplementary Text***

**Comorbidity relations among the whole set of 781 diseases**

For examining whether pre-selecting a subset of diseases with prevalence > 0.1% will affect the comorbidity relations among common diseases or not, we further calculate the comorbidity relations by focusing on the whole set of diseases (781 diseases). We find 12,895 comorbidity relations among the whole set of diseases, in which 9,995 are comorbidities among common diseases (Additional file 2: Table S13). The number of common disease comorbidities is reduced when using the whole set of diseases compared with that when using a subset of diseases (9,995<11,285), because the threshold of *P* values is decreased. Since the ICD10 codes that correspond to the same disease are merged into one disease by phecode^1^, thus the common diseases (prevalence>0.1%) are slightly different when considering the whole set and a subset of diseases. For example, J41, J42, J44 are merged into one disease when considering whole set of disease, but only J42, J44 are merged when considering a subset of diseases. This is because the J41 have prevalence < 0.1% and is removed when only considering diseases with prevalence > 0.1%. There are 373 diseases are commonly used. We find: (1) all the comorbidities (7,762) obtained by using the whole set of diseases are repeated by using a subset of diseases, and (2) 94.5% (7,762/8,218) comorbidities obtained by using a subset of diseases are reproducible when using whole set of diseases. Therefore, the comparison result shows that no matter whether the whole set of diseases or only common diseases (prevalence > 0.1%) are considered, the comorbidity connections among common diseases are almost the same.

**Directionality of the comorbid disease-pairs**

We calculated the directionality between comorbid disease-pairs by the method used by Jensen *et al*^2^. However, we find few comorbidity relations are directional (1,034 comorbidities are directional, and 10,251 are not). This may because the relatively small sample size of UKB hospital inpatient data results in losing power to detect directionality of comorbidity. Since few directional comorbidities are observed, we choose to use both the directional and unidirectional comorbidities for our main analysis, while keeping the directionality of comorbidities available as a reference (Additional file 2: Table S2).

**Comparisons of comorbidities with Hidalgo *et al.* and Jensen *et al.***

Hidalgo *et al.* found 48,038 disease interactions between 995 three digital ICD9 disease codes (RR>1, pval.adj<0.05)^3^. We transform the ICD9 codes into ICD10 codes using the Unified Medical Language System (UMLS)^4^. 148 diseases are commonly used by us and Hidalgo *et al.* Then, we select comorbidities from hidalgo’s results by Bonferroni correction (the correction method used in our article), only 18 disease interactions survive. Therefore, there is no necessary to compare with this small set of comorbidities.

Jensen *et al.* studied the disease trajectories, and found 4,014 directional comorbidities between 681 ICD10 diseases^2^. 384 disease are commonly used by us and Jensen *et al.* We find a significant overlap of comorbidities between the UKB and Jensen’s (OR=6.8, P=0, Fisher exact test). We repeat a relatively high proportion (49%) of Jensen’s comorbidities, compared to 13% of UKB comorbidities repeated by Jensen *et al*. This might be due to the fact that Jensen *et al.* only provided the directional comorbidities and ignored the comorbidities without direction. When we only compare the directional comorbidities in UKB to Jensen’s results, the overlap is also significant (OR=8.9, P=2.7e-148), and the proportion of UKB comorbidities repeated by Jensen *et al.* indeed greatly increases (32%), but the proportion of Jensen’s comorbidities repeated by us reduces to 19%. Furthermore, when prefiltering disease-pairs with 0.1% patients diagnosed them at same day, we find that the overlap is still significant (OR=4.5, P=1.4e-72), and the proportion of UKB comorbidities repeated by Jensen et al. is (32%), and the proportion of Jensen’s comorbidities repeated by us is 32%. Studying the directionality of comorbidities requires a large sample size and a long follow-up time. The diagnosis data in UKB has a long follow-up time (25 years, compared to 15 year by Jensen), but a relatively small sample size (about 0.4 million, compared to 6.2 million by Jensen). Despite this, comparison with Jensen’s results confirms the reliability of our comorbidities.

**Comorbidity tendency intra- and inter- categories based on new chapters**

We have further calculated the disease comorbidity tendency intra- and inter- categories based on the disease classification given by Zhou *et al* ^5^. Zhou *et al.* classified 1,797 diseases into 17 distinct new chapters (NC01-NC17) by integrating phenotypic and molecular networks. As the 1,797 diseases are based on ICD9 codes, thus we firstly map the ICD9 codes to ICD10 codes using the UMLS^4^. 1,209 ICD9 diseases are successfully mapped to 224 ICD10 diseases (Additional file 1: Fig. S10A), only covering 3,484 UKB comorbidities—31% of all the identified comorbidities in our analysis. By using the same method as described in main article (Comorbidity tendency intra- and inter-categories in Method), we only find the diseases within NC05 tend to co-occur (Additional file 1: Fig. S10B). NC05 includes 10 diseases (I05, I08, I10, I21;I22, I25, I35, I42, I46, G25, G95), mainly related to cardiovascular diseases. This comorbidity tendency has been identified in our primary analysis based on physiological system-based disease classification. The failure to observe more comorbidity patterns may be due to the incomplete coverage of diseases in new chapter, as we see, only half of the common diseases is covered by the new chapter, and only about 31% of comorbidities are included.
